# Effect of using personal protective equipment during the COVID-19 pandemic on the quality indicators of screening colonoscopies

**DOI:** 10.1101/2021.05.07.21256743

**Authors:** Subin Chirayath, Janak Bahirwani, Parampreet Kaur, Noel Martins, Ronak Modi

## Abstract

**Background:** Coronavirus Disease 2019 (COVID-19) has affected many facets of the practice of medicine including screening colonoscopies.

**Aims:** Our study looks to observe if there has been an effect on the quality of colonoscopies, as indicated by quality measures such as cecal intubation rate (CIR), cecal intubation time (CIT), scope withdrawal time (SWT) and adenoma detection rate (ADR) with the adoption of standard COVID-19 precautions.

**Methods:** We conducted a retrospective chart review to analyze the effects of the COVID-19 pandemic on screening colonoscopies. The study utilized data on CIR, CIT, SWT and ADR from outpatient, non-emergent procedures conducted at 3 endoscopy suites of St Luke’s University Health Network. All inpatient and emergent procedures were excluded.

**Results:** Our study demonstrated that the total number of screening colonoscopies was decreased between 2019 to 2020 (318 in 2019 vs 157 in 2020, p= 0.005). CIT (320±105 seconds in 2019 vs 392±107 seconds in 2020, p=0.001) and SWT (706±232 seconds in 2019 vs 830±241 seconds in 2020, p=0.001) were increased while CIR (98.2% in 2019 vs 96.6% in 2020, p=0.04) was decreased between 2019 and 2020 likely due to PPE introduction. ADR was similar between the two groups (38.23 (12.50-66.66) in 2019 vs 38.18(16.66-66.00) in 2020, p=0.8).

**Conclusion:** Our study showed that quality indices for screening colonoscopies like CIR, CIT, and SWT were negatively impacted during the COVID-19 time period. ADR, however, were similar. Thus, the efficiency of the procedures was affected by the use of PPE but it did not affect the colonoscopy’s clinical benefit.

## Introduction

SARS-COV-2 or Coronavirus Disease 2019 (COVID-19) has affected many facets of the practice of medicine. It has resulted in an alarming amount of hospitalizations. Since January 21 2020, a total of 27.8 million cases have been diagnosed, in addition to 488,000 deaths in the United States alone.^1^ Due to the massive spread of the pandemic, personal protective equipment (PPE) has become a part of daily routine for healthcare workers. A majority of standard PPE worn today include gown, gloves, N95 mask and face shield or some form of eye protection. It has been shown by numerous studies to decrease the rate of new infections as much as 5% over a relatively short period of time, particularly among healthcare workers.^2^ Strict adherence to wearing PPE has been adopted in most hospitals and is also important in the procedural setting as well.

Initially, outpatient procedures were being deferred in order to minimize transmission. However, emergent surgeries and procedures would continue and as COVID -19 became more predominant in medicine, elective procedures would also return a few months later. One of the procedures that has been closely followed is colonoscopies. Delayed diagnosis in cancer screening during the pandemic was a concern but some studies have shown that there is no effect on cancer detection rates over a 10 month period.^3^ Endoscopic procedures are considered aerosol-generating procedures which means there is a risk of transmission of viruses due to aerosolization when the scope is inserted and removed. The risk of aerosolization during lower GI procedures has not been well studied. Soon after the COVID-19 pandemic, the American Gastroenterological Association (AGA) issued recommendations for GI endoscopy personnel. For all GI procedures the AGA recommends the use of N95 (or N99, or PAPR) and recommends against the use of surgical masks only, regardless of COVID-19 status.^4^ The decision to extend the recommendation to lower gastrointestinal procedures is based on evidence of possible aerosolization during colonoscopy especially during the insertion and removal of instruments through the biopsy channel and the uncertain risks associated with evidence of the presence of the viral RNA in fecal channels.

Our study looks to observe if there has been an negative effect on both the amount of screening colonoscopies and the quality measures commonly reported such as cecal intubation rate (CIR), cecal intubation time (CIT), scope withdrawal time (SWT) and adenoma detection rate (ADR) during the pandemic period with the adoption of standard COVID-19 precautions. We hypothesized that the pandemic caused a significant decrease in the number of screening colonoscopies and other outpatient endoscopies. We also hypothesized that since the major change in protocol for a screening colonoscopy involved the use of PPE, that it could possibly account for any possible changes in the quality indicators.

## Methods

We conducted a retrospective chart review to analyze the effects of the COVID-19 pandemic on GI endoscopy procedures. The comparison was made during the first peak around the time when the AGA issued recommendations for GI endoscopy personnel. We compared the number of procedures performed, type of procedures, CIR, CIT, SWT and ADR between mid-May to mid-June (05/16-06/14) of 2019 to the same time period in 2020 (05/18-06/16). The comparison was done for outpatient, non-emergent procedures conducted at 3 endoscopy suites at St Luke’s University Health Network in Bethlehem, PA, USA. The procedures themselves were performed by the same attending physicians each of which have at least 4-6 years of experience. The same endoscopes were used during both time periods. All inpatient and emergent colonoscopies were excluded. Data was obtained by performing a chart review on EPIC electronic health records. SPSS version 26 was used to analyze the data. Missing values were not analyzed in the data. No Bonferroni correction was applied for multiple comparisons. P-values equal to or less than 0.05 were considered statistically significant.

## Results

There were a total of 1609 patients who underwent procedures during the period of mid-May to mid-June 2019 (Pre-COVID) and 1198 patients during the one-month period of mid-May to mid-June 2020 (COVID). The median age of patients undergoing endoscopy procedures was 59 in 2019 and 61 in 2020. 62% of the patients were males in 2019 as compared to 58% in 2020. Number and type of colonoscopy were compared between pre COVID and COVID time. There was a significant decline in colonoscopies with 1024(63.7%) in 2019 to 637 (53.7%) in 2020. (Table 1) Further classification was done for the colonoscopies to see the difference in the screening and diagnostic colonoscopies between the two years. The number of screening colonoscopies were almost half in 2020 compared to 2019, from 972 in pre-COVID to 648 during COVID (p= 0.005).

**Table 1:** A crosstab between Diagnostic/Screening colonoscopies and Pre-COVID/COVID Time.

| Type of Colonoscopy | Pre-COVID 2019<br>N=1024 | COVID 2020<br>N=637 |
| --- | --- | --- |
| Diagnostic | 706(68.9%) | 480(75.4%) |
| Screening | 318(31.1%) | 157(24.6%) |

p-value=0.005(Chi-Square test)

Independent sample t-tests were done to compare CIT and SWT between pre-COVID and COVID periods (Table 2). The mean cecal intubation time in COVID (392±107 seconds) was significantly higher than that of Pre-COVID group ((320±105), p=0.001). Similarly scope withdrawal time in COVID (830±241 seconds) was significantly higher than Pre-COVID group ((706±232), p=0.001) whereas CIR was significantly lower in COVID (96.60%) compared to Pre-COVID time ((98.20%), p=0.04) (Table 3).

**Table 2:**
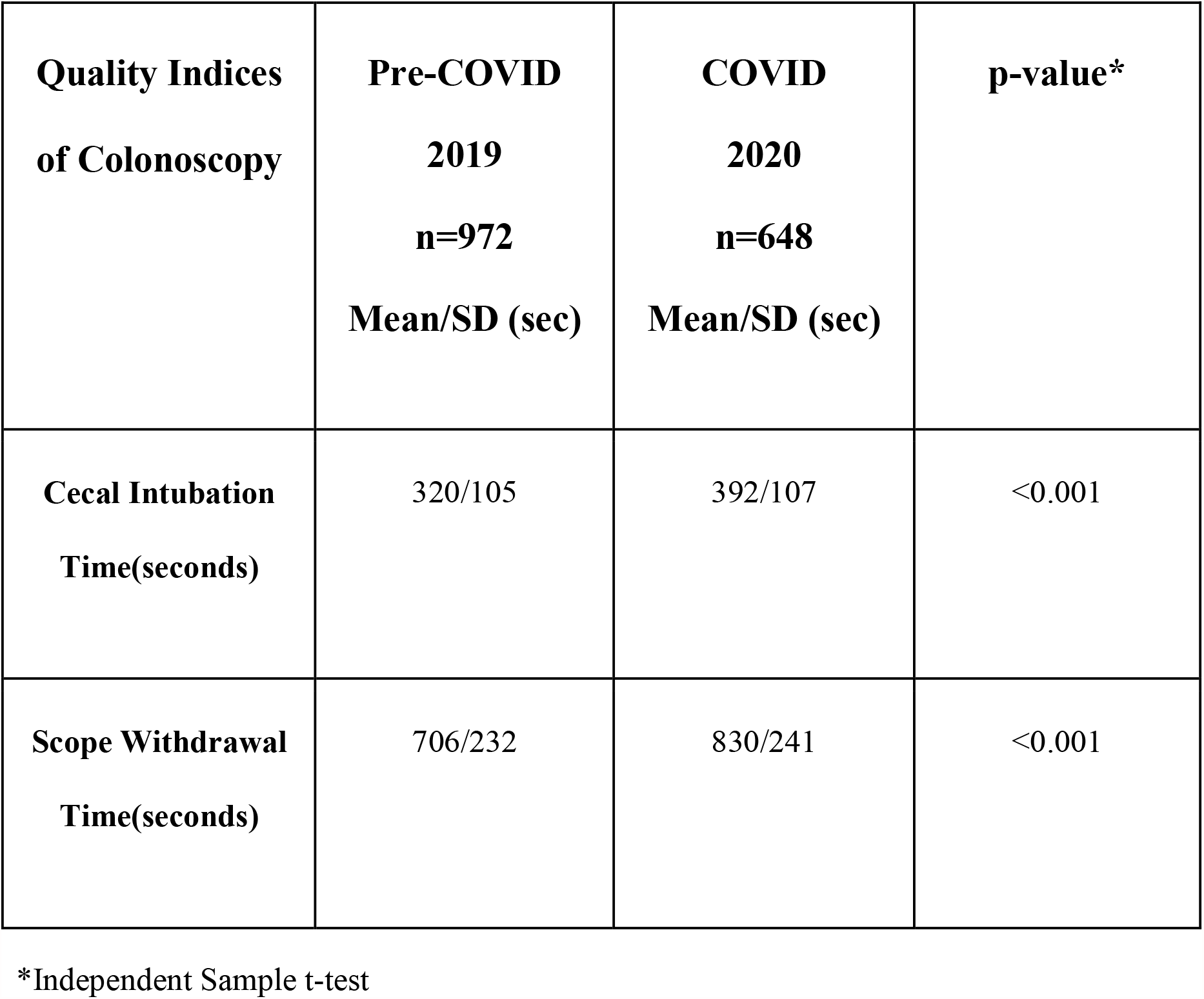
A comparison of cecal intubation times and scope withdrawal times in screening colonoscopies between Pre-COVID-19 (2019) and COVID-19 (2020) time periods.

| **Quality Indices**<br>**of Colonoscopy** | **Pre-COVID**<br>**2019**<br>**n=972**<br>**Mean/SD (sec)** | **COVID**<br>**2020**<br>**n=648**<br>**Mean/SD (sec)** | **p-value\*** |
| --- | --- | --- | --- |
| **Cecal Intubation**<br>**Time(seconds)** | 320/105 | 392/107 | <0.001 |
| **Scope Withdrawal**<br>**Time(seconds)** | 706/232 | 830/241 | <0.001 |

\*Independent Sample t-test

**Table 3:**
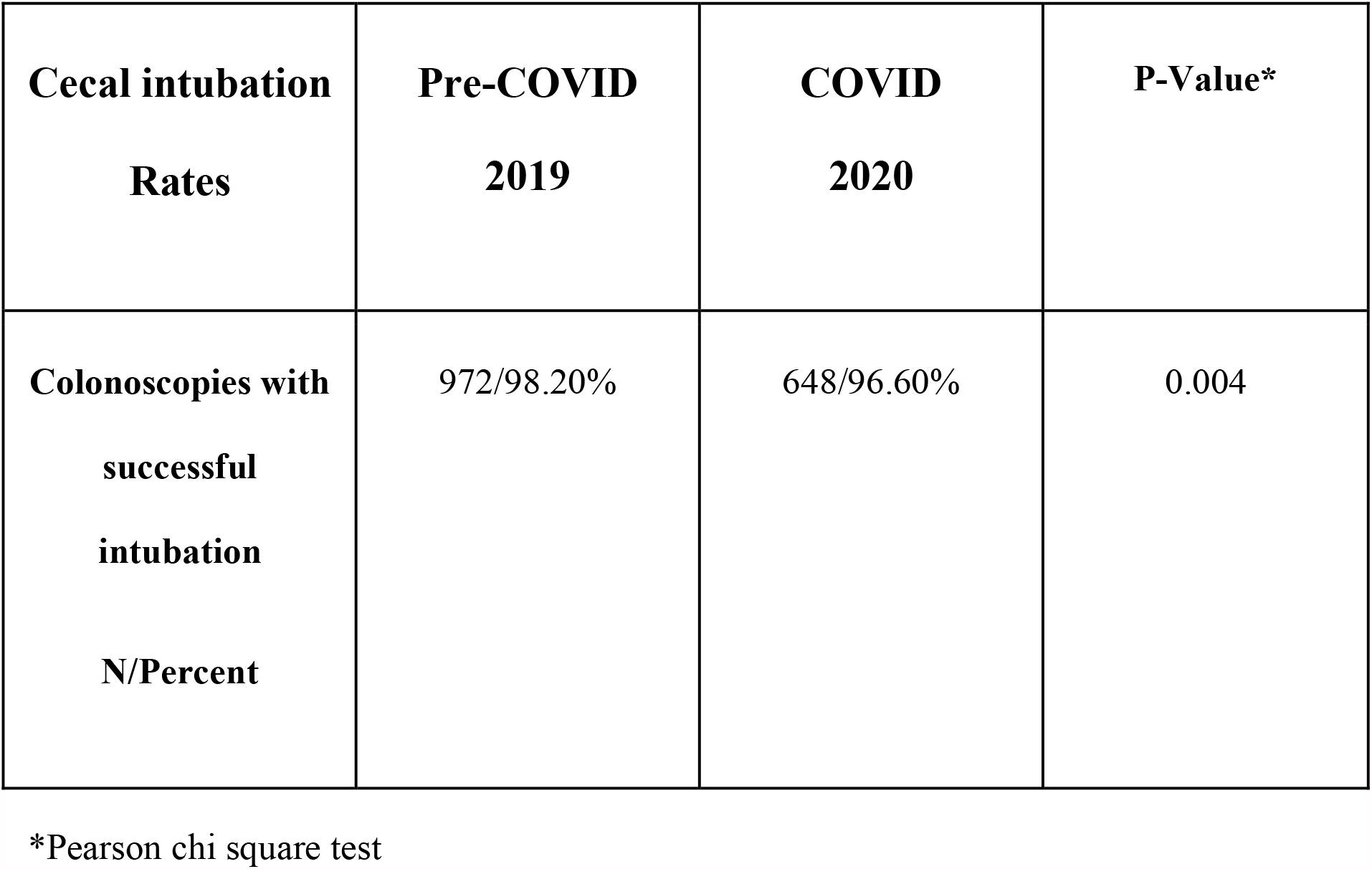
A comparison of cecal intubation rates in screening colonoscopies between Pre-COVID and COVID time

| **Cecal intubation**<br>**Rates** | **Pre-COVID**<br>**2019** | **COVID**<br>**2020** | **P-Value\*** |
| --- | --- | --- | --- |
| **Colonoscopies with**<br>**successful**<br>**intubation**<br>**N/Percent** | 972/98.20% | 648/96.60% | 0.004 |

\*Pearson chi square test

Since the data for adenoma detection rates was not normally distributed, we conducted non-parametric Mann Whitney tests to compare the rates during these two periods (Table 4). It was seen that the median rate of detection during 2019 was 38.23% (12.50-66.66) and during 2020 it was 38.18% (16.66-66.00). The adenoma detection rates between these two periods were not statistically different.

**Table 4:** A comparison of adenoma detection rates between the Pre-COVID-19 (2019) and COVID-19 (2020) time periods.

| **Pre-COVID**<br>**2019**<br>**N=23**<br>**Median**<br>**(min-max)** | **COVID**<br>**2020**<br>**N=22**<br>**Median**<br>**(min-max)** | **P-Value\*** |
| --- | --- | --- |
| 38.23%<br>(12.50-66.66) | 38.18 %<br>(16.66-60.00) | 0.8 |

\*Mann Whitney test

## Discussion

The impact of the global COVID-19 pandemic permeates every facet of medicine, including endoscopic procedures. Our study focused to see if implementing PPE protocols into the routine procedure of a colonoscopy could possibly affect quality indicators. Colonoscopies have been shown to play a vital role in preventing colorectal cancer (CRC). CRC screening during the pandemic overall decreased during the “lockdown period” in early 2020 due to a fear of exacerbating the spread of the virus. Modes of transmission have been studied which include aerosolization of the virus during the procedure as well as fecal contact spread.^5^ Studies however have shown that screening colonoscopies are both safe and efficacious when performed using proper PPE and decontamination protocols for the endoscopic room after every procedure during the pandemic.^6^ The likely causes of the decrease in screening colonoscopies in our study include fear on part of the patient to participate during the pandemic as well as physicians deferring screening colonoscopies and only electing for urgent/diagnostic procedures to minimize the risk of exposure.

Our study demonstrated that CIT and SWT were increased while CIR was decreased between 2019 and 2020 possibly due to PPE introduction. Before the pandemic, many factors have been shown to affect cecal intubation rates and cecal intubation time. CIR is an important quality measure that gastroenterologists are evaluated on. One study showed that age of patient >60, constipation, poor preparation and two person colonoscopies were all independent risk factors for increased cecal intubation rates.^7^ The efficiency of the procedures may be affected due to the standard precautions taken to ensure low transmission of the virus. COVID-19 precautions include thorough cleaning of the room, donning of PPE (which includes N95 mask, gown, gloves, face shield) for all staff during the procedures, and repeating this during turnover for every colonoscopy. This time likely translates to increased procedure time and possible decreased efficiency. This may be an independent risk factor that has not been accounted for in previous studies.

There have been few studies on this subject but one similar one showed no difference in overall procedure time (including cecal intubation rate) between pre and post COVID-19 colonoscopy standards.^8^ This was the opposite of our findings, however, this study had a lower power (N=256) compared to our study which may have skewed their results. Our study demonstrated that CIR was decreased from 2019 to 2020 (p=0.04). It is difficult to say if the implementation of PPE had a direct impact on CIR as this variable is also dependent on other factors including patient anatomy. Overall, PPE may have a negative impact on CIR and CIT. This decreased efficiency however may not have any clinical significance and PPE continues to be necessary during the pandemic to maintain the safety of the practitioners and patients.

Another major aspect of our study focused on the comparison between ADR in colonoscopies prior to the implementation PPE to those performed after the pandemic began. ADR is distinguished from polyp detection rates (PDR) in that the former is a subset of the latter. Some studies have attempted to provide a conversion factor between PDR to ADR.^9^ ADR has been observed to be a valuable marker for cancer related mortality. One study that reviewed over 300,000 colonoscopies found that ADR was inversely related to risk of developing interval advanced stage and fatal colorectal cancer.^10^ In another prospective cohort study, increased ADR was associated with a decrease in cancer related mortality.^11^ We theorized that the additional PPE used during the procedure might obscure a practitioner’s ability to see additional adenomas and thus affect ADR. Surprisingly there was no difference between both groups in our results. Teh et al as discussed prior conducted a similar study which showed no difference in ADR between pre and post COVID-19 precaution colonoscopies. This is reassuring as we see that PPE may not interfere with the clear benefits of colonoscopies.

SWT was also prolonged during colonoscopies done during the pandemic with PPE. SWT is an important measure of the efficacy of a colonoscopy. It acts as a “second pass” to detect lesions in the colon not visible on entry. Interestingly, longer SWT are associated with an increased polyp detection rate, particularly when the time is > 6 minutes.^12^ One study showed that a SWT of 10 minutes was shown to have a higher detection of overall polyps but no difference in detection of adenomatous polyps.^13^ Longer SWT allow for practitioners to be more diligent in the visualization of the entire colon to the end of the procedure. In our study, SWT pre-COVID was approximately 11 minutes, which was closer to the literature values, while during the COVID time period SWT was around approximately 13 minutes. It may not have any clinical significance though this may affect the amount of time a patient is under anesthesia which may lead to associated complications. Our study also found that colonoscopies performed during COVID-19 had prolonged SWT without improving ADR.

Our study did have some limitations. We acknowledge that there are confounding factors including variability between endoscopists in both expertise and experience. However, the same endoscopists performed all colonoscopies that were used between the two time periods to limit variations. The increase in CIT and SWT may or may not have a clinical relevance that was observed in this study. Increased CIT and SWT may translate into more time under anesthesia and could increase complications, though our study did not focus on this. CIR and SWT are also heavily dependent on other factors as discussed prior and PPE may or may not be directly implicated in a direct change in values. PPE may be an independent risk factor resulting in prolonged CIT and SWT while decreasing CIR without increasing ADR. Prospective studies will need to be performed to assess this, especially if COVID-19 becomes endemic and precautions continue to be required in the future.

COVID-19 has profoundly permeated every element of medicine over the past year. Colonoscopies are one of the procedures affected by the pandemic. It is vital in the prevention of CRC and the use of PPE minimizes transmission during the procedure. Our retrospective study conveyed there was an increased CIR and SWT. However, adenoma detection rates were similar, indicating that the use of PPE does not affect a colonoscopy’s efficacy. This did not support our original hypothesis that there may be a negative impact on all quality indicators. With the advent of COVID vaccines, these precautions may change in the near future. We were encouraged to see that PPE does not interfere with clinical benefits of colonoscopies including ADR.

## Conclusion

Our study showed that quality indices for screening colonoscopies like cecal intubation rate, cecal intubation time and scope withdrawal time were negatively impacted during the initial COVID time period compared to pre-COVID time. The study also displayed that though there was a significant decline in both screening and diagnostic colonoscopies during pandemic, adenoma detection rates were comparable. Thus, the efficiency of the procedures was affected by the use of PPE but it did not affect the colonoscopy’s clinical benefit.

## Data Availability

This data is available on figshare.

https://doi.org/10.6084/m9.figshare.14546928.v1

## Abbreviations

ADR: Adenoma detection rate
CIR: Cecal intubation rate
CIT: Cecal intubation time
COVID-19: Coronavirus Disease 2019
CRC: Colorectal Cancer
PPE: Personal protective equipment
SWT: Scope withdrawal time

## Notes

### Competing Interest Statement

The authors have declared no competing interest.

### Funding Statement

This study was not funded by a grant.

### Author Declarations

This study was reviewed and approved by the institutional review board of St. Luke's University Hospital. The following is a statement of their approval: This approval is based on the understanding that you will: - Immediately inform the IRB of all patients serious adverse events and any changes in procedures and project status changes that may occur after this review. - Use only reproductions of the enclosed informed consent form displaying the IRB approval stamp. - Agree to comply with FDA, OPRR, and St Luke's Hospital IRB regulations. - Allow the review of research project records by the IRB as requested. St. Luke's University Health Network has a Federal Wide Assurance [FWA 00003557] from OHRP. The Institutional Review Board is registered with OHRP [IRB 00002757] and is in compliance with 45 CFR 46, 21 CFR 50 and 21 CFR 56. To the extent these Federal regulations are in agreement with the ICH Guidelines, we are also in GCP compliance.

